# Rapid and early diagnosis of tuberculosis in household contacts of cases with the disease in Havana, Cuba (COMBO.X-TB)

**DOI:** 10.64898/2026.01.23.25341243

**Authors:** Raul Diaz-Rodriguez, Alina Martinez-Rodriguez, Maria Rosarys Martinez-Romero, Gladys Abreu-Suarez, Alexander Gonzalez-Diaz, Ellen M.H. Mitchell

**Author notes:** corresponding author Pedro Kourí Institute of Tropical Medicine (IPK), Havana, Cuba.

## Abstract

Since 2014, Cuba’s National Tuberculosis Control and Elimination Program (PNCET, acronym in Spanish) has been using the Xpert® MTB/RIF technique (Cepheid, Sunnyvale, CA, USA) for rapid molecular diagnosis of tuberculosis (TB). Due to financial limitations and difficult access to kits and cartridges (both of which are of U.S. origin), this test is only intended for prioritized vulnerable groups. Thus, only one third of the cases reported annually are diagnosed by rapid tests, as an initial technique.

The scope of Xpert diagnosis can be increased by using the ‘pooling sputum’ method, where several sputum samples from different persons are pooled and applied in a single Xpert® MTB/RIF Ultra (or simply Xpert Ultra) cartridge. This method of pooling sputum with the Xpert Ultra has been used with samples from symptomatic respiratory patients from countries with a high burden of disease, and its use could be extended to household contacts without expectoration, over 8 years of age.

Sputum may not always be produced spontaneously in people with incipient, asymptomatic disease, and children often have problems expectorating. The use of a positive expiratory pressure (PEP) device, Lung Flute (LF) ECO (Acoustic Innovation, Tokyo, Japan), helps to produce sputum more easily and its result could be very similar to the use of a hypertonic solution.

To increase rapid molecular diagnosis in people with suspected TB and at an early stage of the disease, this project aims to: 1. evaluate the pooled sputum method and the Xpert Ultra molecular assay in the rapid diagnosis of TB in household contacts of cases with bacteriologically confirmed pulmonary TB. 2. Estimate the proportion of household contacts that are able to produce a sputum sample suitable for Xpert Ultra using the LF ECO device.

In Latin America, Xpert® MTB/RIF, and more recently Xpert Ultra, is used in almost all countries as the first TB diagnostic technique, but the percentages are still low, mainly due to the scarce economic resources available in the region. Therefore, the use of novel strategies, such as the use of sample mixtures pooled in a single Xpert Ultra cartridge, will increase the coverage of testing for TB cases, reduce the sources of infection in the community and reduce the costs of cases diagnosed by rapid tests.

This study will provide important information on the combination of a powerful molecular tool (Xpert® MTB/RIF Ultra) for TB diagnosis, the application of a pooling sputum method and the use of a PEP device, Lung Flute ECO, in household contacts with and without respiratory symptoms.

The synergistic methods in this study could serve as a reference for other countries in the process of TB elimination. These countries need to vastly increase their testing to find a rapidly shrinking number of cases, and our efficient strategy represents a potentially sustainable way to do this. Low income countries face challenging hurdles to acquire sufficient imported proprietary consumables for high volume testing. Pooling is increasingly viewed as a way to promote equity and access in settings without preferential pricing or favorable trade relations.

## Introduction

Tuberculosis (TB) remains one of the most serious and important health problems worldwide. The World Health Organization (WHO) estimated that 10 million cases and 1.4 million deaths occurred in 2019. Of the estimated total cases, 12% were children <18 years of age, 8.2% were people living with HIV, and nearly half a million had TB resistant to at least rifampicin or with multiple drug resistance [1].

Cuba is one of the 15 countries with the lowest burden of disease in the Americas region [2] and is working steadily to eliminate TB [3]. In 2019, WHO estimated a total incidence rate of 6.5 TB cases per 100 000 population, with 90% being bacteriologically confirmed pulmonary TB [2]. The rate in children/young people under 18 years of age was 1.0% and the estimated proportion of new cases with rifampicin-resistant and multidrug-resistant TB (RR/MDR TB) was only 2.2% [2]. However, the incidence rate has remained around 6 per 100 000 population from 2004 to the present [4].

Since 2014, Cuba’s National Tuberculosis Control and Elimination Program (PNCET) has been applying Xpert® MTB/RIF (Xpert) for rapid TB diagnosis [5]. Due to financial limitations and difficult access to kits and cartridges (both of which are made in the United States), this test is only intended for prioritized vulnerable groups (household or primary contacts of a case of pulmonary TB, people living with HIV/AIDS, persons deprived of their liberty, persons suspected of having resistant TB, foreigners or Cuban aid workers from countries with a high burden of the disease, and children). Thus, this rapid test has been used as an initial diagnostic technique in only about one third of the diagnosed cases in recent years [2].

The scope of Xpert diagnosis of individuals with a high suspicion of TB can be increased, without greatly increasing the costs of the test, if the pooling sputum method is applied [6, 7]. With this method, several sputum samples from different patients (usually three-four samples) can be pooled and the resulting mixture is applied in a highly sensitive Xpert cartridge (Xpert® MTB/RIF Ultra or simply Xpert Ultra). If *Mycobacterium tuberculosis complex* is detected in the pooled sample*, it* means that at least one of the samples is positive. Then, each individual sample will be tested to know which one(s) is (are) positive [6, 7]. This pooled sputum method, tested with the Xpert Ultra cartridge, has been applied to people with characteristic symptoms and signs of TB, especially in high burden countries [8]. However, its use could be extended to household contacts without spontaneous expectoration, older than 8 years of age.

Sputum often cannot be produced spontaneously by people with incipient, asymptomatic disease, so the use of a PEP device to assist those who struggle to expectorate facilitates this task and its outcome may be very similar to the use of a hypertonic solution [9]. The use of the Lung Flute™ (Medical Acoustics, LLC, Buffalo, NY, USA) causes an oscillation of expiratory pressure and airflow, which induces a vibration of airway cells, a decrease in airway collapsibility and an acceleration of airflow [10]. This flute has also been used successfully to induce sputum in children/young adults with cystic fibrosis aged 6-18 years [11]. The LF ECO variant (Acoustic Innovation, Tokyo, Japan), is made of paper, which lowers its cost without losing its functions.

In countries with low TB prevalence and an interest in eliminating the disease, it is important to apply novel strategies to increase TB case-finding and minimize the sources of infection in the community. To increase rapid molecular diagnosis (by Xpert) in persons with suspected TB and at an early stage of the disease, we intend to evaluate the pooled sputum method in household contacts of bacteriologically confirmed pulmonary TB cases and to apply a PEP device for persons without expectoration [8, 12].

In Latin America, Xpert is used in almost all countries as the first TB diagnostic technique, but the percentages are still low, mainly due to the scarce economic resources available in the region [2]. Therefore, the use of novel strategies, such as the use of pooled mixtures in a single Xpert Ultra cartridge, will increase the search for TB cases, minimize the sources of infection in the community and reduce the costs of cases diagnosed by rapid tests.

In Havana, the capital of Cuba, historically about a third of the country’s TB cases occur, and because of the high population density and mobility, combating the disease is very complex. For example, in 2019, 201 new TB cases were reported, representing 34% of the total number of cases [4]. Additionally, a study on TB inequalities in children/youth under 18 years of age in Western Cuba (2011-2015), shows the higher figures of TB in the capital of the country and that the control of the disease needs sustained strengthening to move towards elimination [13]. For this reason, Havana could be a good study site to develop a research of this type.

If the project obtains the expected results, the benefits could include:

- An increase in the early diagnosis of TB in Havana, Cuba, in **household contacts** (not symptomatic), 8 years of age and above, of TB cases using a sensitive molecular tool (Xpert Ultra), a sputum-producing device (LF ECO) and the combination of sputum samples.
- a decrease in the estimated cost of the Xpert Ultra molecular assay in TB case finding by using pooled samples.
- a reduced dependency on foreign diagnostic commodities subject to vagaries of shipping and access
- a reduction of the gap between estimated and reported cases of TB and DR-TB in the coming years in Cuba and take steps towards the elimination of the disease.
- The results of this project could also serve as a reference for other countries in the Americas region in the process of TB elimination.

### Research questions

Is it possible to accurately detect TB in household contacts of patients with bacteriologically confirmed pulmonary TB in the absence of symptoms? Is the pooled sputum method for Xpert Ultra sensitive enough to replace testing on individual samples? Could high quality sputum samples (with detectable *M. tuberculosis complex*) be obtained in people without spontaneous expectoration using the LF ECO device?

### Objectives of the study

1. To evaluate the pooled sputum method and the Xpert Ultra molecular assay in the rapid diagnosis of TB in household contacts of bacteriologically confirmed pulmonary TB cases.
2. Estimate the proportion of household contacts that are able to produce a sputum sample suitable for Xpert Ultra using the LF ECO device.

### Design and methods

#### Context of the study

The study will be carried out in Havana, the capital of Cuba, with an estimated population of 2.1 million inhabitants and a density of 2,918.4 inhabitants/Km^2^ [4, 14]. As the capital, Havana is the province with the highest population density and economic importance, where the main sources of the disease are concentrated and its behavior determines, to a large extent, the results of the nation [15). At the end of 2019, the notification rate of new cases and relapses was 9.5 per 100,000 inhabitants, well above the national average (5.2 per 100,000 inhabitants) [4]. Similar to what has occurred in the last five years (Table 1).

**Table 1.**
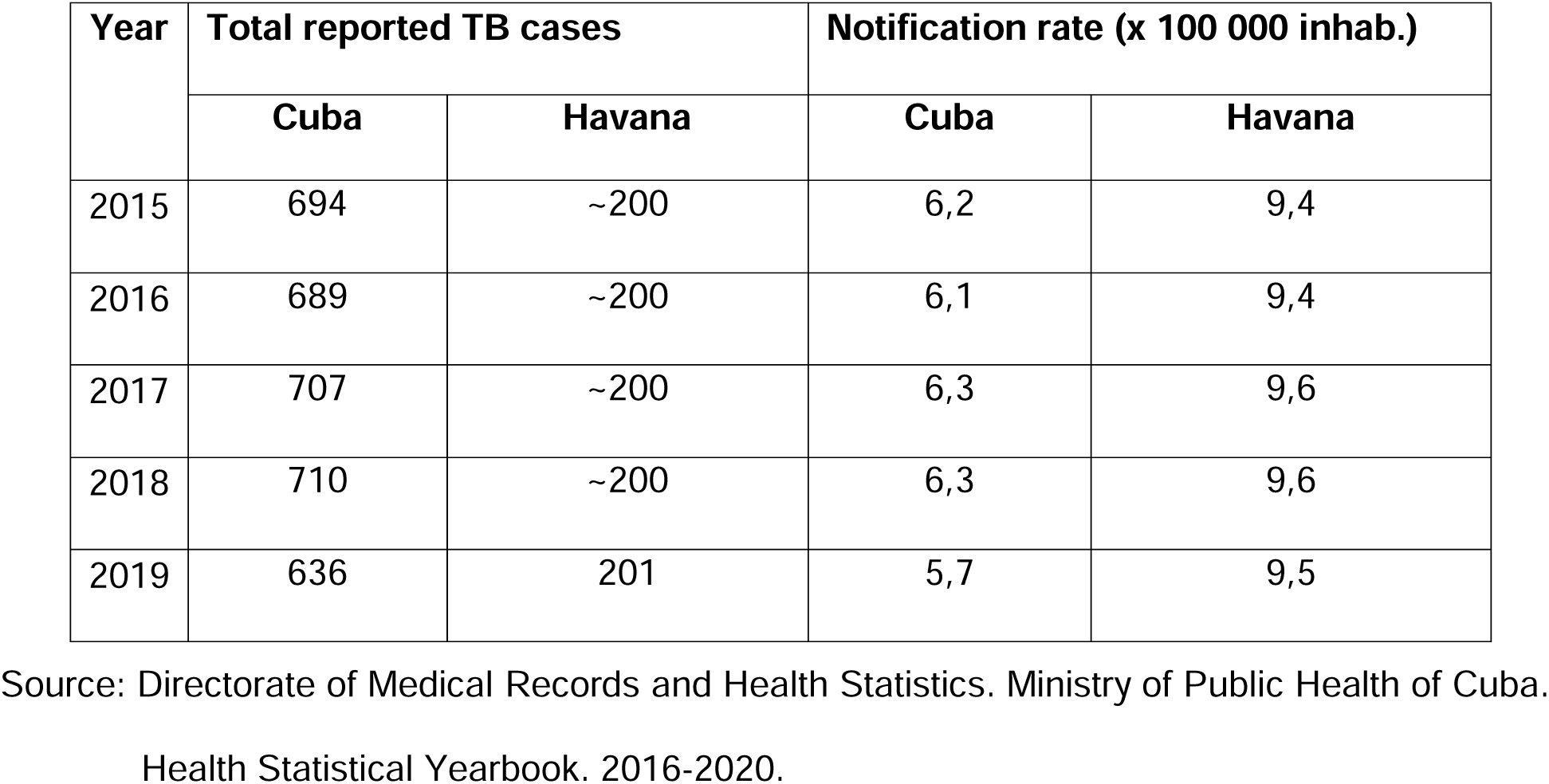
Total notified cases of tuberculosis and notification rate for Cuba and Havana in the last five years.

The Provincial TB Control Program in Havana (and of course the PNCET) bases the search for cases on active screening in prioritized vulnerable groups (VG) (see page 3) and on passive screening in respiratory symptomatic patients (RS) with more than 21 days. In VGs, Xpert MTB/RIF is applied as the first diagnostic test and for the rest of the RS the conventional techniques of microscopy of direct smears for acid-fast bacilli (AFB) and culture in Löwenstein-Jensen (LJ) medium are used [5, 16].

#### Study design

A cross-sectional study will be conducted to evaluate pooled or individual sputum samples for detection of *M. tuberculosis complex* by Xpert-Ultra and the utility of the LF ECO device in active TB case-finding among household contacts 8 years of age and above.

#### Study population (Universe)

Household contacts, 8 years of age and above, of all bacteriologically confirmed pulmonary TB cases notified to the PNCET in Havana between August 2022 and July 2024.

##### Recruitment of contacts

Identification and recruitment of potential study participants will be carried out by PNCET staff in Havana at the provincial, municipal and local levels while routine contact investigation of TB cases is being conducted.

The management of the Provincial Program (included in this project) will systematically inform a principal investigator of the Project about the cases of bacteriologically confirmed pulmonary TB in Havana, and will provide a list of these patients with the names, personal addresses and telephone numbers of all household contacts registered in the routine epidemiological investigation of the PNCET.

Potential participants will be contacted by telephone, preferably, or in person to give their consent to be visited at a later date for the purposes of this research. In-home contacts who agree to a visit to try to participate in the study will be scheduled for a date and time and will be notified at least 72 hours in advance of the proposed visit for acceptance. During the home visit to the potential participant (or to his/her parent or guardian, if he/she is a child/youth under 18 years of age), the objective of the research, the different types of samples to be taken and why, the usefulness of the LF ECO to facilitate expectoration, the diagnostic processes in the laboratory, the risks and inconveniences of taking each sample and the individual and social benefits of participating in this research will be explained in detail (see more in sub-heading Ethical aspects).

### Inclusion criteria

- Household contact of a bacteriologically confirmed pulmonary TB patient.
- Age greater than or equal to 8 years old,
- Consent to participate in the study (in the case of minors, consent must be given by parents or guardians).
- Informed assent (for children/youth 8 to 18 years of age)

### Exclusion and Exit Criteria

#### Exclusion criteria

- Seriously ill persons receiving palliative care at home
- Persons with disabilities that prevent them from providing sputum samples for study (International Classification of Functioning, Disability and Health).

#### Exit criteria

- Intra-domiciliary contacts outside the home during the sampling period.

### Calculation of sample size and cartridges to be used

#### Sample Size Calculation

In the period 2015-2019, about 200 cases were reported annually in Havana (see Table 2). Of these, an average of 180 cases with bacteriologically confirmed pulmonary TB were reported. If it is taken into account that the average number of household contacts of these cases ranged from 8 to 11 people, with a mean of 9, the number of individuals to be sampled should be 1620 (with a range of 1440-1980). As for children/youth under 18 years of age, excluding neonates, who were contacts of bacteriologically confirmed pulmonary TB cases, the mean was 2 with a range between 1-3 children. The total number of children/youth to be sampled should be 360 (ranging from 180 to 540) or 22% of the total contacts in the study. Of the total number of children/youth to be studied, 10-20% may be under 8 years of age. These children may represent 36 and 72 and will not be included in this study due to the potential challenge of mastery of expectoration technique (see section Ethical aspects). If we take into account the lowest percentage (10%) only 36 will be excluded. Therefore, the total sample of household contacts to be studied should be **1584** (with a range of 1426-1944).

**Table 2.**
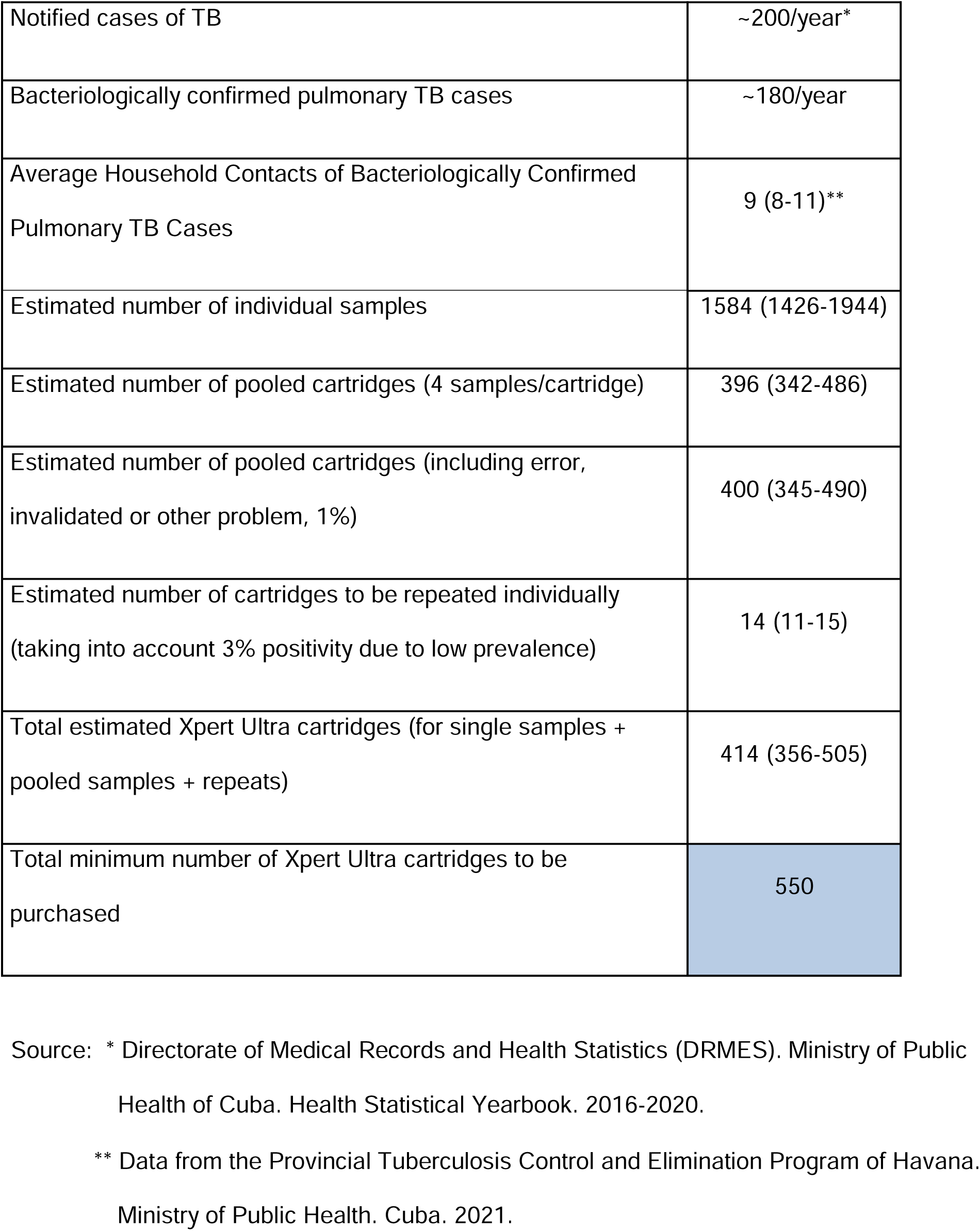
Main Havana TB data (in the last five years) and calculation of samples and Xpert Ultra cartridges to be used.

#### Calculation of total cartridges to be used

##### Sputum samples from all household contacts, older than 8 years of age, will be submitted to the Xpert Ultra technique in a mixture of their pooled sputum (in groups of four sputum)

If all the estimated individuals, 1584 (with a range of 1368-1944), are sampled and divided into pools of 4 people, we would have between 396 pooled cartridges (with a range of 342-486) in total (see Table 2). If we take into account that 1% of these could give results with error, invalid or other problems, the number of cartridges needed to be used would rise to 391-400 (with a range of 345-491). Considering that Cuba is a country with a low prevalence of TB, the expected positivity in these individuals is approximately 3%. Therefore, it will be necessary to repeat test individual samples from a positive pool in no more than 14 individuals (ranging between 11 and 15). If a comprehensive estimate is made of the cartridges used for pooled and individual samples and repeats, the final total would be **414** (range, 356-505). Therefore, the minimum number of cartridges needed should be 550.

## Procedures

### Sampling and Transportation

For a better understanding of the samples and techniques to be used in this protocol see **S4 File**. (Diagnostic Algorithm).

#### Sputum

All household contacts of bacteriologically confirmed pulmonary TB cases, who consent (or their parent or guardian, for children/youth aged 8-18 years) to participate in the investigation, will be asked to provide a sputum sample and will be provided with a sputum collection bottle. Each bottle will be appropriately labeled in accordance with the accompanying specimen order.

##### Sputum produced with Lung Flute (LF) ECO

Contacts who are unable to expectorate naturally will be instructed to use the LF ECO device, which will be kindly provided by the Institute of Tropical Medicine in Antwerp (Belgium) via the DGD FA5 mechanism, following published recommendations [9, 10]. Briefly, after a deep inhalation, the contacts will blow twice into the mouthpiece of the LF ECO (similar to blowing out a lit candle). Then, they will rest for two or more cycles of normal breaths and repeat the process 20 more times, under the care and observation of the research medical staff.

#### Transportation of samples

An IPK investigator and the person in charge of the provincial TB program will be responsible for transporting the samples from the place where they are taken to the national reference laboratory (LNRITBLM, at the IPK). This transport will be carried out in appropriate containers, complying with the established national biosafety standards [17].

### Sample processing and analysis by Xpert® MTB/RIF Ultra (or Xpert Ultra)

#### Sputum

Sputum samples (volume ≥3 mL) will be divided into three parts. 1 mL will be taken for individual analysis, 1 mL for preparing pooled samples and 1 mL for solid culture on LJ medium (reference or "gold" test).

Aliquots of samples should be placed in sterile plastic centrifuge tubes (Falcon type) of 15 mL, previously identified. Samples for individual analysis and for culture should be stored at −20° C until use.

##### Individual sputum samples for Xpert Ultra

Individual samples will only be analyzed by Xpert Ultra if they are included in pools of combined samples where *M. tuberculosis complex* was detected. For this, the usual methodology described [18] will be followed.

##### Pooled sputum samples for Xpert Ultra

Pooled samples will be prepared in small groups, mixing four sputa taken from household contacts, giving priority in each group to contacts of the same TB patient.

One mL of sputum (amount used in the Xpert Ultra test) will be taken from each sample of each group and deposited in a 15 mL plastic centrifuge tube (Falcon type). The resulting mixture is vortexed for 1 minute. Subsequently, from 1 mL of the mixture the recommended protocol will be continued [18]. The rest of the pooled samples that are not used for the Xpert Ultra should be stored at −20° C, in case it is necessary to repeat any of them.

Between each process of preparing pools of pooled samples, the bottom surface of the biosafety cabinet and the tubes containing the pooled samples should be disinfected with 1% chlorine solution. This is followed by wiping with 70% ethanol to reduce corrosion of the chlorine solution on metal surfaces.

##### Individual sputum samples for culture

All individual samples will be processed for culture to detect *M. tuberculosis complex* (and serve as a reference technique). One mL of sputum will be taken and the usual methodology of the national reference laboratory will be followed [16].

### Interpretation of results

Xpert Ultra results will be interpreted as indicated internationally [18].

The pooled samples that give error, invalid, or no results will be repeated to obtain a positive or negative result for the detection of *M. tuberculosis complex*. For this purpose, the remainder of the pooled sample stored in freezer will be used.

Individual specimens that fail, are invalid, or have no results will be repeated to obtain a positive or negative result for the detection of *M. tuberculosis complex*. If necessary, the household contact (or parent or guardian) will be asked for an additional sample, making it clear that it is a new sample.

The results of the pooled Xpert Ultra specimens will not be considered in establishing the criteria of whether a contact has TB or not. Only positive Xpert Ultra results for individual specimens and sputum culture will be used as diagnostic criteria for TB and will be reported immediately to the PNCET.

### Estimation of the proportion of household contacts that are able to produce a sputum sample suitable for Xpert Ultra

#### Evaluation of the quality and quantity of sputum samples

The quantity (in mL) and quality of sputum samples obtained with the LF ECO device will be determined as reported by Sakashita *et al.* [9] and following the modified <u>Miller & Jones</u> [19] classification.

> **S:** Saliva, **M1:** Mucoid without pus, **M2:** Mucoid with suspected pus, **P1:** purulent grade 1 with pus in less than 1/3 of the sample, **P2:** purulent grade 2 with pus in 1/3-2/3 of the sample, **P3:** purulent grade 3 with pus in more than 2/3 of the sample.

At the national TB reference laboratory, trained laboratory technologists will evaluate the macroscopic characteristics of the sputum quality and the volume produced (in mL), compared to a reference bottle, and fill out the corresponding form.

#### Estimation of the proportion of household contacts that are capable of producing a sputum sample suitable for Xpert Ultra

A value of ≥3 mL of sputum will be taken as an adequate amount of sputum in this investigation (remember that at least 3 mL will be needed for individual and pooled analyses by Xpert Ultra, and for culture).

A sample will be considered to be of good quality if it falls in the range of P1-P3. However, no sample will be discarded because of its quality level.

The proportion of household contacts that are capable of producing a sputum sample suitable for Xpert Ultra with (B) and without (A) the LF ECO device and as a whole (C) will be estimated, taking into account:

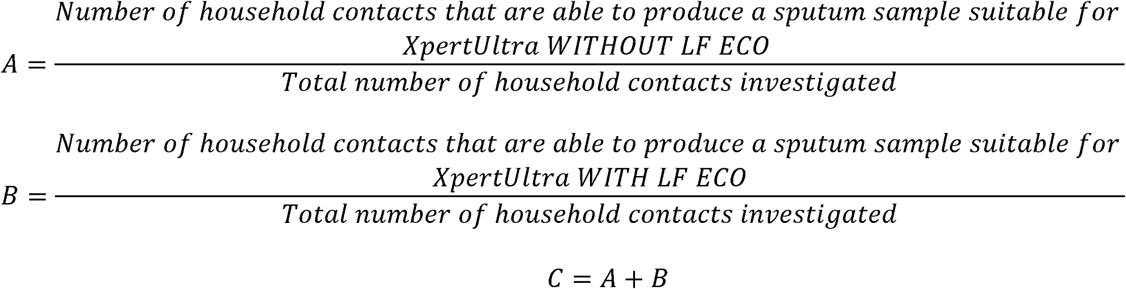

### Estimation of time and resource consumption

#### Estimated time consumed

An estimation of the time (T) consumed for the execution and delivery of Xpert Ultra results will be made, starting from the preparation of the pooled sputum sample (mc) and taking as a reference the time consumed for each individual sample (mi).

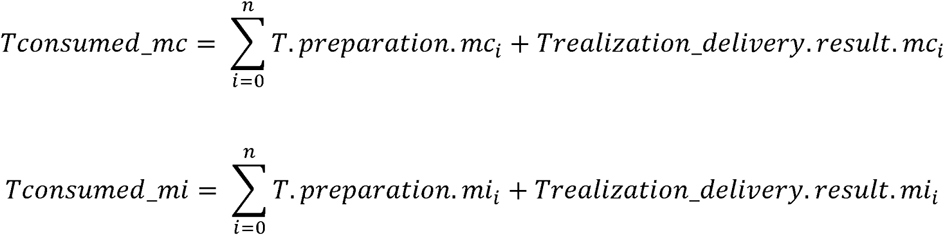

### Estimation of Xpert Ultra Resource Consumption

To estimate the resource consumption of Xpert Ultra, only the cost of Xpert Ultra cartridges will be taken into account, as Xpert Ultra reagents and sterile droppers are included in the diagnostic kit package (there is no additional charge for these).

Number of cartridges used for diagnosis:

- *<u>Number of total cartridges used for diagnosis on pooled samples:</u>* cartridges used for pooled samples + cartridges used on individual samples to define positive pooled sample results + cartridges used to repeat results with error, invalid or other problem.
- *<u>Number of cartridges used for individual samples:</u>* cartridges used for diagnosis on individual samples + cartridges used to repeat results with error, invalid or other problem.

#### Number of Xpert Ultra Cartridges saved

Number of cartridges used for individual negative samples - Number of total cartridges used for diagnosis in negative pooled samples

### Data Management and Statistical Analysis

The data contained in Form 1 (Collection of data on index TB cases) and Form 2 (Investigation of household contacts) in all sections (except for the "fill and review" sections) will be taken from existing primary sources: a) Clinical History from the Family Physician’s Office. b) Epidemiological History (PNCET model 81-51). These data are routinely collected in the contact investigation described in the PNCET [16].

In Form 1, section F, all identified household contacts will be listed; they will be contacted to ask for informed consent to participate in the study. Only those who consent to participate will fill out Form 2. For those who do not agree to participate, only the number of persons by age and sex will be analyzed, as part of the description of the universe to be studied.

Xpert Ultra test results for individual and pooled samples will be collected in a log book (see Annex III) and spreadsheet in Microsoft Excel and a database will be created using the coding scheme recommended by the Global Laboratory Initiative [20].

Data will be reviewed for inconsistencies and will be grouped, organized and processed with the Epi Info program^TM^ 7.2.0.1 (Centers for Disease Control, CDC, Atlanta, GA, USA).

For sputum samples will be calculated:

- <u>Sensitivity (S)</u>: percentage of contacts with *M. tuberculosis complex* detected by solid medium culture and correctly identified by the Xpert test. <u>Specificity (E)</u>: percentage of contacts without identified *M. tuberculosis complex* (by solid medium culture) correctly identified by Xpert Ultra.
- <u>Positive predictive value (PPV)</u>: percentage of contacts with *M. tuberculosis complex* (taking into account the solid medium culture) with a positive Xpert Ultra test.
- <u>Negative predictive value (NPV)</u>: percentage of patients with *M. tuberculosis complex* (on solid culture) with a negative Xpert Ultra test in their specimens.
- <u>Positive likelihood ratio (PLR)</u>: the ratio of the odds that the test will be positive in contacts with *M. tuberculosis* versus those without *M. tuberculosis complex* detected. The higher the PLR (out of 1) the more important the contribution of a positive test result in the diagnosis of the disease.
- <u>Negative likelihood ratio (NLR)</u>: the ratio of the odds of testing negative in contacts with *M. tuberculosis* to those without *M. tuberculosis complex* detected. The RVN values the contribution of a negative result in the "non-confirmation" of disease.
- <u>Youden Index (YI):</u> statistical test that will report the performance of the pooled sample method: YI = Sensitivity + Specificity - 1

In addition, performance analyses of the LF ECO device and the diagnostic capability of the use of Xpert Ultra pooled samples will be performed according to the individual characteristics of the contacts grouped according to sex, age ranges, and other variables of interest.

Data on index TB cases and housing characteristics will allow the exposure of household contacts and the likelihood of infection/disease in them to be assessed.

The surveys applied to the contacts who used the LF ECO will make it possible to evaluate the occurrence of adverse events from the user’s perspective and the acceptance of its use. Percentages will be calculated per response to each question designed. This survey will be validated before its application in the study.

### Ethical aspects

This study received formal approval by the Research Ethics Committee of IPK (see S1 File), the PAHO Ethics Review Committee (PAHOERC) (S2 File), The International Review Board (IRB) of ITM (S3 File), the PNCET and other authorities from the Ministry of Public Health (Minsap) of Cuba.

It is important to note that children under 8 years of age had to be excluded from this investigation (**molecular diagnosis of TB in household contacts**). In this age group, most of the children with high suspicion of TB struggle to master expectoration techniques. We will develop a companion study to explore a novel method that is child-friendly (tongue swabs).

In this study, a single-use paper PEP device, LF ECO, will be used to facilitate expectoration if the individual is unable to expectorate spontaneously. The use of this ‘LF’ is simple and produces few adverse effects and discomfort that disappear quickly without any treatment; however, its use could mean an added risk for the participant. This paper device (LF ECO) is not registered in Cuba; however, a similar flute has been used successfully in patients (children) with cystic fibrosis in this country. For this reason, the supplier Acoustic Innovation (Tokyo, Japan) and the Center for State Control of Medicines, Medical Equipment and Devices (Cuban regulatory body) have already been contacted so that the supplier can send the pertinent information and its use in the country can be authorized. The LF device has been used in pediatric patients in Australia [11] and also in adults [10]. The study will not interfere with the care that participants should routinely receive and the follow-up by health professionals in their area.

Participants will not receive incentives for participating in this study. They will have the direct benefits of rapid diagnosis of the disease (if present) and early access to curative or preventive treatment as required. The results of this research can help redefine and adapt TB control strategies in Cuba and support progress towards elimination of the disease in the country.

The confidentiality of the participants will be ensured through the codification of the participants’ identification data in compliance with principle 24 of the Declaration of Helsinki. The codes generated will be stored in a key-protected file and in an Excel sheet protected with passwords and stored on a PC with no connection to computer networks. The rest of the information will be worked on using the established codes and will also be protected with passwords. The information will be used to communicate to the physician in charge of patient care and by the principal investigator (PI) of this project for the analysis of the results. De-coding will be performed when a positive result is found (establishing the diagnosis of TB, by any of the techniques employed), in order for the patient to access the benefits of programmatic TB case management.

The identified contacts (or their parents or guardians, in the case of minors under 18 years of age) will give their consent to participate. They will be given an informed consent document and will be explained what this research consists of, their participation, and what it would mean for them individually and for the National TB Program. For children older than 11 years, the study, its risks and advantages will be explained to them in simpler words. Then, they will be shown an informed assent document to sign if they agree to participate, making it clear to them that their parents/guardians are aware of this consultation and agree to their involvement. For children between 8 and 11 years of age, a verbal script was prepared that should facilitate understanding of the study to be conducted and help to achieve greater acceptance.

The follow-up of contacts will be as established by the program: those who are TB cases will receive the established treatment and those who are not, will be followed for two years with periodic evaluations and children and in people living with HIV will be offered preventive therapy.

The collection of sputum samples with LF ECO should be performed by trained personnel using personal aerosol protection equipment, following the recommendations of the national biosafety standards [16].

Used LF ECO devices should be disposed of properly following established national protocols [16].

Samples should be transported in appropriate containers, complying with the established national biosafety standards [16].

The work with the samples will be carried out in a class II Biosafety Cabinet (according to national and international classifications) to work with mycobacteria (16]. In this way, the transmission of pathogenic microorganisms to the exterior will be prevented, so that such work does not represent a risk for the community where the laboratories are located. The personnel in charge have sufficient knowledge of biosafety standards for handling samples and performing the techniques to be implemented.

## Discussion

Cuba is one of the leading countries in the fight against TB in the Americas region [2] and is working strongly to end the disease. In this sense, introduce new techniques and methods to speed up the diagnosis, treatment and prevention is crucial.

This study will provide important information on the combination of one of the most powerful molecular tools (Xpert® MTB/RIF Ultra) for TB diagnosis, the application of a pooling sputum method (to reduce costs) and the use of a PEP device, Lung Flute ECO, in household contacts with and without respiratory symptoms.

Main expected results: 1. Increased coverage of rapid diagnosis of TB from Havana, Cuba, in household contacts (non-respiratory symptomatic) of TB cases using a sensitive molecular tool (Xpert Ultra) and a sputum-producing device (LF ECO) in children/youth aged 8-18 years and adults, excluding young children and neonates. 2. Decrease in the estimated cost of the Xpert Ultra molecular assay in TB case finding by using pooled samples. 3. Reducing the gap between estimated and reported cases of TB and DR-TB in the coming years and taking giant strides toward elimination of the disease.

The synergistic methods in this study could serve as a reference for other countries in the process of TB elimination. Countries with a low TB burden need to vastly increase their testing to find a rapidly shrinking number of cases, and our efficient strategy represents a potentially sustainable way to do this. Low income countries face challenging hurdles to acquire sufficient imported proprietary consumables for high volume testing. Pooling is increasingly viewed as a way to promote equity and access in settings without preferential pricing or favorable trade relations.

### Limitations of the study

One of the main limitations of the study is that the experimental period will be only two years, mainly due to the unavailability of additional financial resources.

Working with people who tend not to manifest any respiratory signs and symptoms of having TB will lower the probability of expectoration and perceived level of importance given by potential participants which could lead to increased refusal rate in household contacts.

Experience in the use of the LF ECO device in the facilitation of expectoration for TB is lacking in the Cuban population and this could lead to low acceptability among health workers.

The simultaneous execution of the project tasks and the unpredictability of the COVID-19 epidemic (or other major health problem) in Cuba, or particular situations of the PNCET, could slow down its development and even halt it in some health areas.

### Quality assurance

#### Quality of Information

Principal Investigator will be responsible for data extraction, supervision of data collection, data analysis and reporting, in accordance with the protocol; which will be reviewed and supervised by another person (project manager) and errors detected, if applicable, will be corrected and documented with date and signed by both parties. The primary data and other files related to the study will be shared with all parties involved to ensure adequate assistance and to collaborate with the quality control of the research process, including tabulation and analysis of the data obtained.

#### Data management and archiving

All data and observations will be entered into a log book and Excel spreadsheets, kept in a secure location. Participant data will be de-identified as soon as it is linked and entered into a tablet using Open Data Kit. After completion of the study, all relevant documentation will be preserved for a period of ten years, in accordance with international requirements.

### Communication and dissemination of results

The results of this research will be reported upon completion of the study in publications in peer-reviewed journals (national and international journals) and will be presented at national and international conferences and workshops. Communication and publication of the results of the study will be done jointly by researchers from IPK, ITM, and other Minsap‘s participants.

## Data Availability

All data produced in the present study will be available upon reasonable request to the authors

## Acknowledgments

We would like to thank the reviewers of the local (IPK, ITM) and international (PAHO Ethics Review Committee, PAHOERC) ethics committees and the consultants from the local PAHO Office in Havana and from PAHO Headquarters (Washington) for their assistance.

## Author Contributions

Conceptualization: Raul Diaz-Rodriguez, Alina Martinez-Rodriguez, Ellen M.H. Mitchell.

Funding acquisition and project administration: Raul Diaz-Rodriguez, Ellen M.H. Mitchell.

Methodology: Raul Diaz-Rodriguez, Alina Martinez-Rodriguez, Maria Rosarys Martinez-Romero, Gladys Abreu-Suarez, Ellen M.H. Mitchell.

Formal analysis: Raul Diaz-Rodriguez, Alina Martinez-Rodriguez, Ellen M.H. Mitchell.

Writing-original draft: Raul Diaz-Rodriguez, Alina Martinez-Rodriguez, Maria Rosarys Martinez-Romero, Ellen M.H. Mitchell.

Writing-review & editing: Raul Diaz-Rodriguez, Alina Martinez-Rodriguez, Maria Rosarys Martinez-Romero, Gladys Abreu-Suarez, Alexander Gonzalez-Diaz, Ellen M.H. Mitchell.

## Supporting information

**S1 File.** Letter of communication of the approved protocol by the Research Ethics Committee (CEI), Pedro Kouri Institute of Tropical Medicine (IPK), Havana, Cuba. (PDF)

**S2 File.** Letter of communication of the approved protocol by the PAHO Ethics Review Committee (PAHOERC), Pan-American Health Organization (PAHO). (PDF)

**S3 File.** Letter of communication of the approved protocol by the Institutional Review Board (IRB) of Institute of Tropical Medicine (ITM), Antwerp, Belgium. (PDF)

**S4 File.** Diagnostic algorithm of Tuberculosis according to this protocol. (PDF)

