## Supplemental Document 1 for "Rapid and early diagnosis of tuberculosis in household contacts of cases with the disease in Havana, Cuba (COMBO.X-TB)"

**RESEARCH PROTOCOL CEI-IPK 27-21**

### 'Impact of rapid and early diagnosis of contacts of tuberculosis cases (including children) in the disease elimination stage in Cuba'


**PRINCIPAL INVESTIGATOR**

**PhD. Raúl Díaz-Rodríguez**


After the evaluation and analysis of this document by the members of the institution's Ethics Committee, following the international working guidelines of these committees of the World Health Organization, we issue the following:

#### OPINION

1. The document presented conforms to the principles established by the Declaration of Helsinki as well as to the ethical standards and criteria established in the national codes of ethics and legal regulations in force in Cuba.
2. The protocol clearly reflects the ethical aspects that are appropriate for the type of research proposed.
3. **APPROVED**, the submitted document.

Given at the IPK, Havana, Cuba, on the 29th day of November 2021

DrCs. Eric Martínez Torres
Presidente CEI-IPK

**NOTE: This is an English translation of the offcial letter (in Spanish) from CEI-IPK**
