## Supplemental Document 2 for "Rapid and early diagnosis of tuberculosis in household contacts of cases with the disease in Havana, Cuba (COMBO.X-TB)"

**Pan-American Health Organization**
**Ethics Review Committee (PAHOERC)**

Evaluation Report for Research Proposals

**PAHOERC Ref. No: PAHOERC.0462.03**

| | |
|---|---|
| Unit: | CDE |
| Proposal title: | **Impact of rapid and early diagnosis of contacts of tuberculosis cases in the elimination stage of the disease in Cuba** |
| Principal Investigator: | Raul Diaz Rodriguez |
| Focal Point:: | Freddy Perez |
| Country(es): | Cuba |

PAHOERC reviewed this project and concluded on May 16, 2022 that it has been **approved** for implementation in Cuba.

------------------------------

We encourage researchers to plan for the publication and use of their research results. We promote publication in open-access, indexed journals.

Please inform local review committees of this decision. You must inform PAHOERC if there are significant changes in the implementation of this proposal.

Carolina Chávez

Carolina Chavez Cortes, Member, PAHOERC 16-may-22

Carla Saenz, Secretary, PAHOERC 16-may-22

**Note: A copy of this letter, approved and signed by PAHOERC, must be included in the documentation attached when requesting the preparation of contracts, letters of agreement, or legal documents from PAHO relevant to research projects.** **05/16/2022**

**IMPORTANT NOTE: This is an English translation of the offcial letter (in Spanish) from PAHOERC**
