## Supplemental Document 3 for "Rapid and early diagnosis of tuberculosis in household contacts of cases with the disease in Havana, Cuba (COMBO.X-TB)"

Research Protocol, Version 3.2, date: May 4, 2022

| | |
|---|---|
| **Sponsor:** | TDR/OPS |
| **Responsible for the Project in Cuba:** | Raúl Díaz Rodríguez |
| **Department/Laboratory:** | Bacteriology-Mycology. National Reference Laboratory for Tuberculosis, Leprosy and Mycobacteria. |
| **Complete mailing address:** | **Instituto de Medicina Tropical "Pedro Kourí" (IPK)**, Autopista Novia del Mediodía Km 6 1/2, P.O. Box 601, La Lisa, Havana 11400 |
| **Telephone numbers:** | *Telephone (professional):* (537)255-3526/3527/3528<br>*Telephone (mobile/cellular):* (535)470-6847 |
| **Email:** | *Main e-mail:*<br>*Secondary e-mail:* |
| # **Protocol:** | *Not applicable* |

| | |
|---|---|
| **Coordinating investigator at ITM** | *Ellen M.H.Mitchell (DPH)* |
| **Title:** | *Rapid and early diagnosis of tuberculosis in household contacts of cases with the disease in Havana, Cuba (COMBO.X-TB)* |
| **Version:** | *3.2 (May 4, 2022)* |

| | |
|---|---|
| **Main Institution:** | - Pedro Kourí Institute of Tropical Medicine (IPK), Havana, Cuba:<br>o National Reference and Research Laboratory for Tuberculosis, Leprosy and Mycobacteria (LNRITBLM):<br>o Tuberculosis, Acute Respiratory Infections and Leprosy Research and Surveillance Group. |
| **Other Participating Institutions:** | - Provincial Center of Hygiene, Epidemiology and Microbiology (CPHEM) Havana:<br>- Centro Habana Pediatric University Hospital (HPCH)<br>- National Directorate of Epidemiology (DNE). Ministry of Public Health (Minsap)<br>• Institute for Tropical Medicine, Antwerp, Belgium |
| **Principal Investigators:** | Raul Diaz Rodriguez, Alina Martinez Rodriguez, Maria Rosarys Martinez Romero, Gladys Abreu Suarez, Alexander Gonzalez Diaz, Ellen M.H. Mitchell |

### DECLARATION OF CONFORMITY AND CONFIDENTIALITY

The information contained in this study protocol is privileged and confidential. As such, it may not be disclosed unless specific permission is given in writing by the IPK or when such disclosure is required by national or other laws or regulations. These restrictions on disclosure shall apply equally to all future information provided that is privileged or confidential.

Once the final protocol has been issued and signed by the Investigator(s) and authorized signatories, it cannot be altered informally. Amendments to the protocol have the same legal status and must go through the mandatory review and approval stages before being implemented.

By signing this document, the Investigator(s) agree(s) to conduct the study in accordance with the protocol, applicable ethical guidelines such as the Declaration of Helsinki and in accordance with international scientific standards, as well as all applicable regulatory requirements. The Investigator(s) will also make every reasonable effort to complete the study within the designated timelines.

**PRINCIPAL INVESTIGATOR:** DrC. Raúl Díaz Rodríguez. Senior Researcher. Full Professor. Head of the National Reference and Research Laboratory for Tuberculosis, Leprosy and Mycobacteria (LNRITBLM). Department of Bacteriology-Mycology. Research, Diagnostic and Reference Center. IPK

Date: 04-05-2022

Signature:

Fecha: 04-05-2022

Firma: 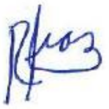

Ellen M.H. Mitchell 7-6-22

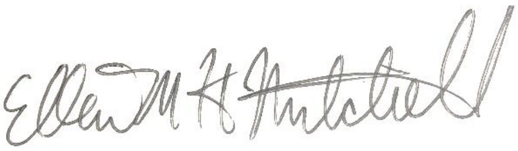

### SUMMARY

Since 2014, Cuba's National Tuberculosis Control and Elimination Program (PNCET) has been using the Xpert® MTB/RIF technique (Cepheid, Sunnyvale, CA, USA) for rapid molecular diagnosis of tuberculosis (TB). Due to financial limitations and difficult access to kits (only four in the country) and cartridges (both of which are of U.S. origin), this test is only intended for prioritized vulnerable groups. Thus, only one third of the cases reported annually are diagnosed by rapid tests, as an initial technique.

The scope of Xpert diagnosis can be increased by using the *pooling sputum* method, where several sputum samples from different persons are pooled and applied in a single Xpert® MTB/RIF Ultra (or Xpert Ultra) cartridge. This method of pooling sputum with the Xpert Ultra has been used with samples from symptomatic respiratory patients from countries with a high burden of disease, and its use could be extended to intra-household contacts without expectoration, over 8 years of age.

Sputum may not always be produced spontaneously in people with incipient, asymptomatic disease, and children often have problems expectorating. The use of a positive expiratory pressure (PEP) device, such as *Lung Flute* ECO (Acoustic Innovation, Tokyo, Japan), helps to produce sputum more easily and its result could be very similar to the use of a hypertonic solution.

To increase rapid molecular diagnosis (by Xpert Ultra) in people with suspected TB and at an early stage of the disease, this project aims to:

*Objectives*

1. -To evaluate the pooled sputum method and the Xpert Ultra molecular assay in the rapid diagnosis of TB in household contacts of cases with bacteriologically confirmed pulmonary TB.
2. Estimate the proportion of household contacts that are able to produce a sputum sample suitable for Xpert Ultra using the *Lung Flute* ECO device.

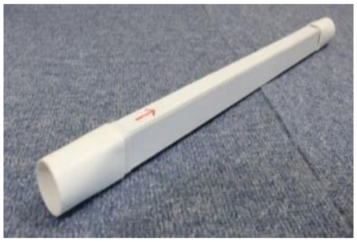

In countries with low TB prevalence and an interest in eliminating the disease, it is important to apply novel strategies to increase TB case-finding and minimize the sources of infection in the community. To increase rapid molecular diagnosis (by Xpert) in persons with suspected TB and at an early stage of the disease, we intend to evaluate the pooled sputum method in household contacts of bacteriologically confirmed pulmonary TB cases and to apply a PEP device for persons without expectoration (8, 12).

**Table 1- Total notified cases of tuberculosis and notification rate for Cuba and Havana in the last five years.**

| Year | Total reported TB cases | | Notification rate (x 100 000 inhab.) | |
|---|---|---|---|---|
| | Cuba | Havana | Cuba | Havana |
| 2015 | 694 | ~200 | 6,2 | 9,4 |
| 2016 | 689 | ~200 | 6,1 | 9,4 |
| 2017 | 707 | ~200 | 6,3 | 9,6 |
| 2018 | 710 | ~200 | 6,3 | 9,6 |
| 2019 | 636 | 201 | 5,7 | 9,5 |

Source: Directorate of Medical Records and Health Statistics. Ministry of Public Health of Cuba. Health Statistical Yearbook. 2016-2020.

The Provincial TB Control Program in Havana (and of course the PNCET) bases the search for cases on active screening in prioritized vulnerable groups (VG) (see sub-section I.1, page 7) and on passive screening in respiratory symptomatic patients (RS) with more than 21 days. In VGs, Xpert MTB/RIF is applied as the first diagnostic test and for the rest of the RS the conventional techniques of bacilloscopy and culture in Löwenstein-Jensen (LJ) medium are used (5, 16).

**III.2. Study design:**

A cross-sectional study will be conducted to evaluate pooled or individual sputum samples for detection of *M. tuberculosis Complex* by Xpert-Ultra and the utility of the *Lung Flute* ECO device in active TB case-finding among intra-household contacts 8 years of age and above.

III.6.2 Calculation of total cartridges to be used

Sputum samples from all intra-household contacts, older than 8 years of age, will be submitted to the Xpert Ultra technique in a mixture of their pooled sputum (in groups of four sputum).

If all the estimated individuals, 1584 (with a range of 1368-1944), are sampled and divided into pools of 4 people, we would have between 396 pooled cartridges (with a range of 342-486) in total (see Table 2). If we take into account that 1% of these could give results with error, invalid or other problems, the number of cartridges needed to be used would rise to 391-400 (with a range of 345-491). Considering that Cuba is a country with a low prevalence of TB, the expected positivity in these individuals is approximately 3%. Therefore, it will be necessary to repeat test individual samples from a positive pool in no more than 14 individuals (ranging between 11 and 15). If a comprehensive estimate is made of the cartridges used for pooled and individual samples and repeats, the final total would be **414** (range, 356-505). Therefore, the minimum number of cartridges needed should be 550.

**Table 2- Main Havana TB data (in the last five years) and calculation of samples and Xpert Ultra cartridges to be used.**

| | |
|---|---|
| Notified cases of TB | ~200/year*<br>~200/year*<br>~200/year*<br>~200/year*<br>~200/year |
| Bacteriologically confirmed pulmonary TB cases | ~180/year |
| Average Household Contacts of Bacteriologically Confirmed Bacteriologically Confirmed Pulmonary TB Cases | 9 (8-11)** |
| Estimated number of individual samples | 1584 (1426-1944) |
| Estimated number of pooled cartridges (4 samples/cartridge) | 396 (342-486) |
| Estimated number of pooled cartridges (including error, invalidated or other problem, 1%) | 400 (345-490) |
| Estimated number of cartridges to be repeated individually (taking into account 3% positivity due to low prevalence) | 14 (11-15) |
| Total estimated Xpert Ultra cartridges (for single samples + pooled samples + repeats) | 414 (356-505) |
| Total minimum number of Xpert Ultra cartridges to be purchased | 550 |

Source: * Directorate of Medical Records and Health Statistics (DRMES). Ministry of Public Health of Cuba. Anuario Estadístico de Salud. 2016-2020.
** Data from the Provincial Tuberculosis Control and Elimination Program. Ministry of Public Health. Cuba. 2021.

### III.7 Procedures

#### III.7.1 Sampling and Transportation

For a better understanding of the samples and techniques to be used in this protocol see Annex I.3- Diagnostic Algorithms.

III.7.1.1 Sputum

All intra-household contacts of bacteriologically confirmed pulmonary TB cases, who consent (or their parent or guardian, for children/youth aged 8-18 years) to participate in the investigation, will be asked to provide a sputum sample and will be provided with a sputum collection bottle. Each bottle will be appropriately labeled in accordance with the accompanying specimen order.

III.7.1.1.1.1 Sputum produced with *Lung Flute* ECO

Contacts who are unable to expectorate naturally will be instructed to use the *Lung Flute* (LF) ECO device, which will be kindly provided by the Institute of Tropical Medicine in Antwerp (Belgium) via the DGD FA5 mechanism, following published recommendations (9, 10). Briefly, after a deep inhalation, the contacts will blow twice into the mouthpiece of the LF ECO (similar to blowing out a lit candle). Then, they will rest for two or more cycles of normal breaths and repeat the process 20 more times, under the care and observation of the research medical staff (see Annex I. 4).

III.7.1.2 Transportation of samples

An IPK investigator and the person in charge of the provincial TB program will be responsible for transporting the samples from the place where they are taken to the national reference laboratory (LNRITBLM, at the IPK). This transport will be carried out in appropriate containers, complying with the established national biosafety standards (17).

III.7.2 Sample processing and analysis by Xpert® MTB/RIF Ultra (or Xpert Ultra)

III.7.2.1 Sputum

Sputum samples (volume ≥3 mL) will be divided into three parts. 1 mL will be taken for individual analysis, 1 mL for preparing pooled samples and 1 mL for solid culture on LJ medium (reference or "gold" test).

Aliquots of samples should be placed in sterile plastic centrifuge tubes (Falcon type) of 15 mL, previously identified. Samples for individual analysis and for culture should be stored at -20º C until use.

III.7.2.1.1 Individual sputum samples for Xpert Ultra

At the national TB reference laboratory, trained laboratory technologists will evaluate the macroscopic characteristics of the sputum quality and the volume produced (in mL), compared to a reference bottle, and fill out the corresponding form (see Annex I.5. Form on sputum collection in intra-household contacts).

<u>III.8.2- Estimation of the proportion of household contacts that are capable of producing a sputum sample suitable for Xpert Ultra.</u>

A value of ≥3 mL of sputum will be taken as an adequate amount of sputum in this investigation (remember that at least 3 mL will be needed for individual and pooled analyses by Xpert Ultra, and for culture).

A sample will be considered to be of good quality if it falls in the range of P1-P3. However, no sample will be discarded because of its quality level.

The proportion of household contacts that are capable of producing a sputum sample suitable for Xpert Ultra with (B) and without (A) the LF ECO device and as a whole (C) will be estimated, taking into account:

$$A = \frac{\begin{array}{c}\text{Número de contactos intradomiciliarios capaces de producir una muestra de esputo adecuada}\\ \text{para XpertUltra SIN LF ECO}\end{array}}{\text{Número total de contactos intradomiciliarios investigados}}$$

- number of household contacts that are able to produce a sputum sample suitable for Xpert Ultra without LF ECO / total number of household contacts investigated

$$B = \frac{\begin{array}{c}\text{Número de contactos intradomiciliarios capaces de producir una muestra de esputo adecuada}\\ \text{para XpertUltra CON LF ECO}\end{array}}{\text{Número total de contactos intradomiciliarios investigados}}$$

- number of household contacts that are able to produce a sputum sample suitable for Xpert Ultra with LF ECO / total number of household contacts investigated

$$C = A + B$$

- number of household contacts that are able to produce a sputum sample suitable for Xpert Ultra (with and without LF ECO) / total number of household contacts investigated

### III.9. Estimation of time and resource consumption

<u>III.9.1 Estimated time consumed</u>

An estimation of the time (T) consumed for the execution and delivery of Xpert Ultra results will be made, starting from the preparation of the pooled sputum sample (mc) and taking as a reference the time consumed for each individual sample (mi).

$$Tconsumido\_mc = \sum_{i=0}^{n} T.preparación.mc_i + Trealiz\_entrega.result.mc_i$$

$$Tconsumido\_mi = \sum_{i=0}^{n} T.preparación.mi_i + Trealiz\_entrega.result.mi_i$$

III.9.2 Estimation of Xpert Ultra Resource Consumption

To estimate the resource consumption of Xpert Ultra, only the cost of Xpert Ultra cartridges will be taken into account, as Xpert Ultra reagents and sterile droppers are included in the diagnostic kit package (there is no additional charge for these).

Number of cartridges used for diagnosis:

- *Number of total cartridges used for diagnosis on pooled samples:* cartridges used for pooled samples + cartridges used on individual samples to define positive pooled sample results + cartridges used to repeat results with error, invalid or other problem.

- *Number of cartridges used for individual samples:* cartridges used for diagnosis on individual samples + cartridges used to repeat results with error, invalid or other problem.

***Number of Xpert Ultra Cartridges saved:*** Number of cartridges used for individual negative samples - Number of total cartridges used for diagnosis in negative pooled samples

**III.10 Data Management and Statistical Analysis**

The data contained in Form 1 (see Annex I.8, Collection of data on index TB cases) and Form 2 (Investigation of intra-household contacts) in all sections (except for the "fill and review" sections) will be taken from existing primary sources: a) Clinical History from the Family Physician's Office. b) Epidemiological History (PNCET model 81-51). These data are routinely collected in the contact investigation described in the PNCET (16).

In Form 1 (Annex I.8), section F, all identified intra-household contacts will be listed; they will be contacted to ask for informed consent to participate in the study (see Annexes I.1 and I.2). Only those who consent to participate will fill out Form 2. For those who do not agree to participate, only the number of persons by age and sex will be analyzed, as part of the description of the universe to be studied.

### IV. ETHICAL ASPECTS

This study received formal approval from the IPK Ethics Committee and is in the process of review by the PAHO Ethics Commission (PAHOERC). The objectives and procedures of the study should be submitted to the PNCET and Minsap authorities for approval.

It is important to note that children under 8 years of age had to be excluded from this investigation (**molecular diagnosis of TB in intra-household contacts**). In this age group, most of the children with high suspicion of TB struggle to master expectoration techniques. We will develop a companion study to explore a novel method that is child-friendly (tongue swabs).

In this study, a single-use paper positive expiratory pressure (PEP) device, *Lung Flute* ECO, will be used to facilitate expectoration if the individual is unable to expectorate spontaneously. The use of this "Lung Flute" is simple and produces few adverse effects and discomfort that disappear quickly without any treatment; however, its use could mean an added risk for the participant. This paper device (*Lung Flute* ECO) is not registered in Cuba; however, a similar flute has been used successfully in patients (children) with cystic fibrosis in this country. For this reason, the supplier *Acoustic Innovation* (Tokyo, Japan) and the Centro Estatal de Control de Equipos Médicos y Dispositivos de Salud, CECMED (Cuban regulatory body) have already been contacted so that the supplier can send the pertinent information and its use in the country can be authorized. The *Lung Flute* device has been used in pediatric patients in Australia (11) and also in adults (10). The study will not interfere with the care that participants should routinely receive and the follow-up by health professionals in their area.

### VII. BUDGET AND SCHEDULE

**VII.1. Budget:** US$18,784.50 (TDR/OPS) Euro €34, 856 (FA5 DGD)

#### VII.2. Schedule

| **YEAR** | **2022** | | | | **2023** | | | | **2024** | | | |
|---|---|---|---|---|---|---|---|---|---|---|---|---|
| **Quarters** | **1** | | | | **1** | | | | **1** | | | |
| Approval by the Ethics Committees | | | X | X | | | | | | | | |
| Evaluation of Xpert Ultra and LF ECO | | | | X | X | X | X | X | X | X | | |
| Data analysis and interpretation | | | | | | | | | | | X | |
| Dissemination of results | | | | | | | | | | | | X |

### VIII. REFERENCES

WHO. Global Tuberculosis Report 2020. Geneva: World Health Organization; 2019. Licence: CC BY-NC-SA 3.0 IGO.

Tuberculosis in the Americas. Regional report 2019. Washington, D.C.: Pan American Health Organization; 2020. License: CC BY-NC-SA 3.0 IGO; https://iris.paho.org/handle/10665.2/52815.

González Ochoa E, Díaz Rodríguez R, Suárez Alvarez L, Abreu Suárez G, Armas Pérez L, Beldarraín Chaple E, et al. Elimination of tuberculosis in Cuba: recent contributions, results and challenges. Rev Cubana Med Trop. 2017;69(3). [accessed: 19/02/2021]. Available at: http://scielo.sld.cu/pdf/mtr/v69n3/a10_260.pdf

4. Directorate of Medical Records and Health Statistics. Ministry of Public Health of Cuba. Health Statistical Yearbook. 2019. 2020 [accessed: 19/02/2021]. Available at: http://files.sld.cu/bvscuba/files/2020/05/Anuario-Electr%C3%B3nico-Espa%C3%B1ol-2019-ed-2020.pdf

Martínez-Romero MR, Secretário-Chilemo T, Lemus-Molina D, Mederos-Cuervo LM, Sardiñas-Aragón M, García-León G, Echemendía-Font M, Díaz-Rodríguez R. Evaluation of Xpert MTB RIF for tuberculosis diagnosis and rifampicin resistance detection in vulnerable groups. Neumol Cir Thorax. 2019;78(3): 284-9.

6. Abdurrahman, S.T.; Mbanaso, O.; Lawson, L.; Oladimeji, O.; Blakiston, M.; Obasanya, J.; Dacombe, R.; Adams, E.R.; Emenyonu, N.; Sahu, S.; et al. Testing pooled sputum with Xpert MTB/RIF for diagnosis of pulmonary tuberculosis to increase affordability in low-income countries. J Clin Microbiol. 2015; 53: 2502-8.

7. Zishiri, V.; Chihota, V.; McCarthy, K.; Charalambous, S.; Churchyard, G.J.; Hoffmann, C.J. Pooling sputum from multiple individuals for Xpert® MTB/RIF testing: A strategy for screening high-risk populations. Int J Tuberc Lung Dis. 2015, 19, 87-90.

8. Chry M, Smelyanskaya M, Ky M, Codlin AJ, Cazabon D, Eang MT, Creswell J. Can the high sensitivity of Xpert MTB/RIF Ultra be harnessed to save cartridge costs? Results from a pooled sputum evaluation in Cambodia. Trop Med Infect Dis. 2020, 5, 27: 2-10.

9. Sakashita K, Fujita A, Takamori M, Takayuki N, Matsumoto T, Saito T. et al. Efficiency of the Lung Flute for sputum induction in patients with presumed pulmonary tuberculosis. Clin Respir J. 2017;00:1-7.

Medical Acoustics. Product overview. Lung Flute® operation. [Accessed 17 February 2021]. Available at: http://www.medicalacoustics.com/Home/LungFlute/Overview/LungFluteOperation

11. Doumit M, Jaffé A. Use of the Lung Flute for sputum induction in children with cystic fibrosis: a pilot study. Pediatr Pulmonol. 2015;50(4):340-3. (see attached bibliography).

12. Ho,J, Jelfs P, Nguyen PTB, Sintchenko V, Fox G J, Marks GB. Pooling sputum samples to improve the feasibility of Xpert MTB/RIF in systematic screening for tuberculosis. Int J Tuberc Lung Dis. 2017;21(5):503-8.

13. Ferrán Torres RM, Abreu Suárez G, González Ochoa E. Inequalities of tuberculosis in children under 15 years of age in Western Cuba, 2011-2015. Rev Cubana Ped. 2021;93(1):e1302.

National Office of Statistics and Information (ONEI). Population and Housing Census. Cuba. 2012. [accessed: 03/02/2021]. Available at: http://www.onei.gob.cu/node/13001

González Díaz A, Sánchez Valdés L, Armas Pérez L et al. Trends and prognosis of tuberculosis, challenges in the elimination stage in Havana. Rev Cuba Med Trop. 2012; 64:163-75.

16. Ministry of Public Health. Ministerial Resolution 277/2014. National Tuberculosis Control Program. Manual of norms and procedures. Havana: Editorial Ciencias Médicas 2014 [accessed 13/09/2021]. Available at: http://www.sld.cu/galerias/pdf/sitios/tuberculosis/programa_2015.pdf

17. Official Gazette of the Republic of Cuba. Resolution 199: Regulations on Biological Safety for the use of biological agents and their products, organisms and fragments of these with genetic information. Havana: Official Gazette of the Republic of Cuba; 2020 [accessed 13/09/2021]. Available at: https://www.gacetaoficial.gob.cu/es/resolucion-199-de-2020-deministerio-de-ciencia-tecnologia-y-medio-ambiente

18. World Health Organization, 2014. Xpert MTB/RIF Implementation manual. Technical and operational 'how-to': practical considerations. France. WHO/HTM/TB/2014.1.

19. Miller DL. A study of techniques for the examination of sputum in a field survey of chronic bronchitis. Am Rev Respir Dis. 1963; 88: 473-83.

20. Global Laboratory Initiative. Practical guide to implementing a Quality Assurance System for Xpert MTB/RIF Testing ("Xpert QA Guide"). 2019. [accessed: 03/02/2021]. Available from: http://www.stoptb.org/wg/gli/assets/documents/Xpert-QA-guide-2019.pdf.

### IX. LIST OF ABBREVIATIONS AND ACRONYMS

-GV : Vulnerable Groups
-IP : Principal Investigator
-IPK : Institute of Tropical Medicine "Pedro Kourí".
-LJ : Löwenstein-Jensen
-LNRITBLM : National Reference and Research Laboratory for Tuberculosis, Leprosy and Mycobacteria.
-MDR : multidrug resistant
-WHO : World Health Organization
-PAHO: Pan American Health Organization
-PNCET : National Program for the Control and Elimination of Tuberculosis.
-TB : Tuberculosis
- pTB: pulmonary tuberculosis
-HIV : human immunodeficiency virus

### X. ANNEXES

#### ANNEX I. INFORMED CONSENT AND ASSENT AND INSTRUCTIONS

#### ANNEX I.1. INFORMED CONSENT DOCUMENT FOR ADULTS (see Annex document)

#### ANNEX I.2. INFORMED CONSENT DOCUMENT FOR CHILDREN/YOUNG PEOPLE AGED 8-18 YEARS (see Annex document)

#### ANNEX I.2.1. INFORMED CONSENT DOCUMENT FOR CHILDREN/YOUNG PEOPLE AGED 12-18 YEARS (see Annex document)

#### ANNEX I.2.2. INFORMED CONSENT DOCUMENT FOR CHILDREN AGED 8-11 YEARS (see Annex)

#### ANNEX I.3. DIAGNOSTIC ALGORITHMS (see Annex document)

#### ANNEX I. 4. INSTRUCTIONS FOR USE OF THE *LUNG FLUTE* ECO DEVICE (see attached document)

#### ANNEX I. FORM ON THE COLLECTION OF SPUTO IN HOUSEHOLD CONTACTS (see Annex document)

#### ANNEX II. EQUIPMENT, MATERIALS AND SUPPLIES AND REAGENTS TO BE USED IN THE PROTOCOL (see Annex document)

#### ANNEX III. LOGBOOKS AND DATABASES FOR XPERT ULTRA (see Annex document)
