## Supplementary figures and images for "Rapid and early diagnosis of tuberculosis in household contacts of cases with the disease in Havana, Cuba (COMBO.X-TB)"

### Supplemental Document 4

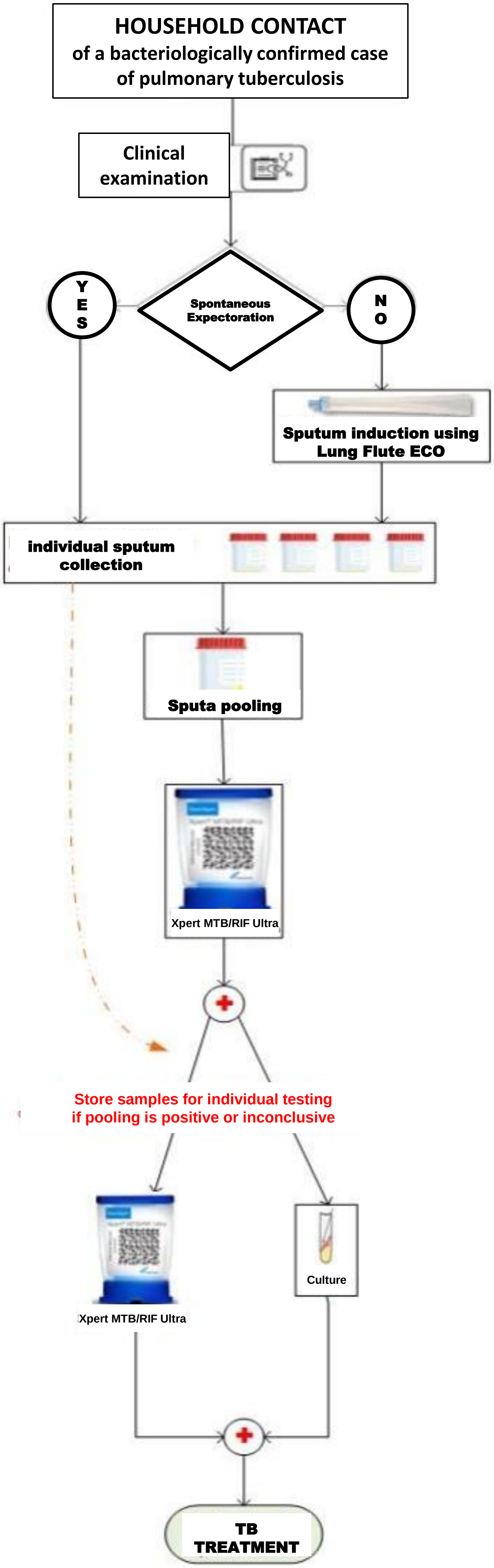
